# Adult mortality in children exposed to state care: systematic review and meta-analysis of prospective cohort studies

**DOI:** 10.1101/2021.09.20.21263839

**Authors:** G. David Batty, Mika Kivimäki, Philipp Frank

**Affiliations:** Department of Epidemiology and Public Health, University College London, London, UK

## Abstract

**Background:** Removal from family of origin to placement in state care is a highly challenging and increasingly prevalent childhood experience. The purpose of this report was to synthesise published and unpublished prospective evidence on adult mortality in people with a history of state care in early life.

**Methods:** For this systematic review and meta-analysis, we searched PubMed and Embase from their inception to May 31^st^ 2021, extracting standard estimates of association and variance from qualifying studies. We augmented these findings with analyses of unpublished individual-participant data from the 1958 and 1970 Birth Cohort Studies (total N = 21,936). Study-specific estimates were aggregated using random-effect meta-analysis. The Cochrane Risk of Bias Tool was used to assess study quality. This review is PROSPERO-registered (CRD42021254665).

**Findings:** We identified 209 potentially eligible published articles, of which 11 prospective cohort studies from the UK, Sweden, Finland, the USA, and Canada met the inclusion criteria (2 unpublished). In 2,273,998 individuals (10 studies), relative to those without a care history in childhood, those who were exposed had 2.5 times the risk of total mortality in adulthood (summary rate ratio; 95% confidence interval: 2.58; 1.96 to 3.39), study-specific estimates varying between 1.53 and 5.77 (I^2^=92%). Despite some attenuation, this association held following adjustment for other measures of early life adversity; extended into middle- and older-age; was stronger in higher quality studies; and was of equal magnitude according to sex and geographical region. There was a suggestion of sensitive periods of exposure to care, whereby individuals who entered public care for the first time in adolescence (3.54; 2.00 to 6.29) experienced greater rates of total mortality than those doing so earlier in the life course (1.69; 1.35 to 2.12). In five studies capturing 1,524,761 individuals (5 studies), children in care had more than three times the risk of competed suicide in adulthood (3.37; 2.64 to 4.30) with study-specific estimates ranging between 2.42 and 5.85 (I^2^=68%). The magnitude of this relationship was weaker after adjustment for multiple covariates; in men versus women; and in lower quality studies.

**Interpretation:** The excess rates of total and suicide mortality in children exposed to state care suggest child protection systems and social policy following care graduation are insufficient to mitigate the effects of the adverse experiences of care itself and the social disadvantage that preceded it.

**Funding:** None.

**Research in context:** *Evidence before this study:* Exposure to state care during childhood has emerging links with an array of unfavourable social, psychological, and behavioural characteristics in early adulthood. We searched PubMed and Embase from their inception to May 31^st^ 2021 for studies examining whether care is also related to elevated rates of adult mortality. While we identified a series of relevant studies, there was no synthesis of this evidence. Few studies utilised a prospective design such that the assessment of care was made in childhood, so avoiding biases of distant retrospective recall. There was also a lack of clarity regarding: the role of confounding factors; the influence of the timing of care entry on mortality; whether the impact of care extended into middle-age and beyond; and, as has been hypothesised, if men with a care history have a greater vulnerability than women.

*Added value of this study:* We conducted a systematic review to synthesis evidence on adult mortality risk in children placed in state care. Drawing also on unpublished resources to complement the findings of published studies, a total of 10 studies consistently showed that exposure to state care in childhood was associated with more than a doubling in the risk of total mortality. This association, while attenuated, held following statistical adjustment for other early life risk factors, including other adversities; extended into later adulthood such that it did not exclusively occur immediately following graduation from care; was stronger in better designed studies; and was of equal magnitude in men and women. There was also a suggestion of sensitive periods of exposure to care, whereby individuals who entered public care for the first time in adolescence experienced greater rates of total mortality in adulthood than those doing so earlier in the life course. The magnitude of the association between childhood care and adult risk of completed suicide (5 studies) were somewhat higher than for total mortality. This relationship was not completely explained by control for other early life risk factors; and the magnitude was somewhat weaker in lower quality studies, and in men versus women. There were too few studies to explore the impact of care on other causes of mortality.

*Implications of all the available evidence:* In recent years there has been a secular rise in the prevalence of children in state care in western societies. This excess mortality risk in this group did not appear to be attributable to other measures of adversity, suggesting that, in the countries studied, child protection systems and social policy following care graduation are insufficient to mitigate the effects of the adverse experiences of care itself and the unfavourable events that preceded it.

## Introduction

Early life is regarded as a critical period of neurobiological, psychological, social, and physical development in all animal species.^1 2^ Via these and other pathways, there is growing evidence that pre-adult adverse experiences may have long-term implications for health across the life course.^3^ Adversity in this context comprises material deprivation^4^ but also stressful family dynamics (e.g., familial psychiatric illness, including alcoholism) and loss or the threat thereof (e.g., parental separation, including incarceration). Of these, removal from family of origin to placement in the care of the state is a particularly challenging early life experience.^5^

Children in state care – also known as out-of-home care, public care, being looked-after, or substitute care – have been temporarily or permanently transferred to alternative accommodation owing to unfavourable events in their home environment, the inability of the biological parents to provide a safe milieu, and/or their own anti-social behaviour, amongst other reasons.^6^ In recent years there has been a secular rise in the prevalence of looked-after children in western societies;^7-9^ while current estimates vary by country and ethnicity, they may be as high as 1 in 10.^10^

Although the decision to transfer to public care is predicated upon providing a better opportunity for the child to flourish, there is growing evidence that such individuals continue to be disadvantaged in several ways. In the short term, relative to their unexposed peers, children in substitute care tend to perform less well academically,^11,12^ are more prone to permanent school exclusion,^13^ are more likely to engage in illicit drug taking,^14^ and have a higher prevalence of a range of psychiatric disorders.^15^ As looked after children transition to independent living in early adulthood, there is a suggestion that these mental health problems continue,^13,16,17^ in addition to there being a greater likelihood of socio-economic disadvantage,^17,18^ homelessness,^19^ and engagement in harmful health behaviours.^17,20^ This burden of risk raises the possibility that exposure to public care in childhood may be associated with an elevated occurrence of adverse physical health outcomes some years following graduation from care, most obviously for mortality. In particular, given the correlation between childhood care exposure and established risk factors for completed suicide – mental health problems,^13^ substance abuse,^21^ and unemployment^22^ – there is a *prima facie* case for care being linked to the intentional taking of one’s own life. While some prospective studies find that care is related to a four-fold elevated risk of total mortality,^23^ others report more modest effects that could be explained by residual confounding.^24^ Results for completed suicide are similarly heterogeneous.^25,26^ To the best of our knowledge, there is no comprehensive and systematic synthesis of the evidence base for the impact of pre-adult care on later total and suicide mortality. Furthermore, the role of the timing of being taken into care is unclear. The notion of ‘sensitive periods’ in this context posits that entering care in adolescence,^27^ as opposed to infancy when the perception of personal circumstances are less acute, may have the most pronounced impact as reflected by an elevated mortality risk.

To address these uncertainties, we conducted a systematic review of all published findings on childhood public care and adult mortality. In doing so, gaps in evidence were, where possible, addressed using individual-participant analyses of unpublished (raw) data from long-term mortality surveillance of participants in two UK birth cohort studies. Lastly, using a meta-analytical approach, we aggregated both sets of results.

## Methods

This systematic review and meta-analysis was registered with PROSPERO (CRD42021254665), a prospectively recorded database.^28^ Reporting of the present results followed the guidelines of Meta-analysis Of Observational Studies in Epidemiology (MOOSE),^29^ Preferred Reporting Items for Systematic Reviews and Meta-Analyses (PRISMA) of diagnostic test accuracy,^30^ and PRISMA of individual participant data.^31^

### Search strategy, study selection, and results extraction for the published studies

We conducted a systematic search of the literature using PubMed (Medline) and Embase databases between their inception in 1966 and May 31^st^ 2021. Without applying any restrictions, we used the terms “out-of-home care”, “out of home care”, “foster care”, “public care”, “looked-after-children”, and “looked after children” for the exposure, and “mortality”, “death”, “suicide”, “cardiovascular”, “stroke”, “heart disease”, and “cancer” for the outcomes. Additionally, we scrutinized the reference sections of retrieved publications for additional reports.

We included a published paper provided it fulfilled all the following criteria: appearing in a peer-reviewed journal; published in English; a prospective study in which the assessment of care was made at ≤18 years of age; mortality surveillance continuing into adulthood; and standard estimates of association (e.g., relative risk, odds ratios, hazard ratios) and variance (e.g., confidence interval, standard error) were provided. When there were multiple reports featuring the same material (e.g., Stockholm birth cohort study^32-34^), the publication with the longest duration of follow-up was included on the basis that this offered the greater statistical power. Retrieved studies were identified by GDB and any potential controversial selections were resolved amongst the authors.

Where available, we extracted and tabulated a range of characteristics from each retrieved paper, including the name of the lead author, publication year, country of sample population, number of participants, year of birth of participants, number of participants exposed to childhood care, number of deaths, and effects estimates for both minimally- and multivariable-adjusted results. Where clarification regarding results or additional analyses were required, we attempted to contact the authors with differing levels of success.

### Individual-participant data for the unpublished studies

We also used individual participant data from two unpublished sources: the 1958 Birth Cohort study (also known as the National Child Development Study) and the 1970 Birth Cohort study. Described in detail elsewhere,^35,36^ these are on-going, closed, geographically-representative, prospective birth cohort studies in which the investigators sampled all live births occurring during a single undisclosed week in 1958 (17,634 babies) and 1970 (17,287 babies) in the contiguous countries that comprise the UK. Data collection for the 1958 cohort study was approved by the National Health Service Research Ethics committee,^35^ and for the 1970 cohort study by the London Central Research Ethics Committee.^36^ With present data being anonymised, additional permissions were not required.

In these studies, state care status was reported by the parents of study members in the first three childhood surveys: ages 7, 11, and 16 years in the 1958 cohort study,^35^ and at ages 5, 10, and 16 years in the 1970 cohort study.^36^ We created a binary variable denoting care in childhood (yes; no). In addition to the standard covariates of sex, parental socioeconomic position (as indexed by occupational social class), and mother’s age at birth of participant, we also adjusted effect estimates for: adverse childhood experience (denoted by physical neglect; child’s or family’s contact with prison service; parental separation due to divorce, death or, other; family history of mental illness; family history of substance abuse); childhood disability; and psychological distress as measured at age 7 in the 1958 cohort study using the teacher-rated Bristol Social Adjustment Guide,^37^ and at age 5 in the 1970 cohort study using the Rutter Behaviour Scale.^38^ Vital status was derived from official death certificates provided using the National Health Service Central Register^39^ and/or fieldwork and cohort maintenance work (<1% of deaths). Members of the 1958 cohort were followed up for 42 years for total (all-cause) mortality from March 1974 (age 16) until December 2016 (age 58), while participants in the 1970 cohort were followed up for 27 years from March 1986 (age 16) until December 2013 (age 43).

### Quality assessment

We used the Cochrane Risk of Bias Tool for cohort studies to assess study quality.^40^ Comprising seven domains, including the exposure, confounding variables, outcome, adequacy of the follow-up and so on, we regarded the quality of the study as high if all were assessed favourably (supplementary table 1).

### Statistical analysis

In analyses of individuals without missing data in the 1958 and 1970 birth cohort studies, we ascertained if the proportional hazards assumption had been violated by computing Schoenfeld residuals.^41^ With no strong evidence that this was the case, we computed hazard ratios with accompanying 95% confidence intervals for care and total mortality separately for each study. Next, we pooled the results from the analyses of raw data alongside published study specific estimates using a random effects meta-analysis; an I^2^ statistic was computed to summarise the heterogeneity in estimates across studies. Lastly, we explored the magnitude of the care–mortality association according to different contexts, including sex, study quality, region, statistical adjustment, and so on. All analyses were computed using Stata 15 (StataCorp, College Station, TX).

### Role of the funding source

This report had no external funding. All authors had final responsibility for the decision to submit for publication.

## Results

We identified 209 published studies of which 9 met our inclusion criteria; a further two were unpublished (**figure 1**). There was a sufficiently large number of studies reporting on the association between care and mortality from all-cause (8 published reports;^10,23,24,26,42-45^ 2 unpublished) and suicide deaths (5 published reports^23-25,42,43^) to facilitate meta-analysis, whereas the frequency of the presentation of other outcomes, such as cancer^42^ and external causes combined,^42,44^ was too low to be included herein.

**Figure 1.**
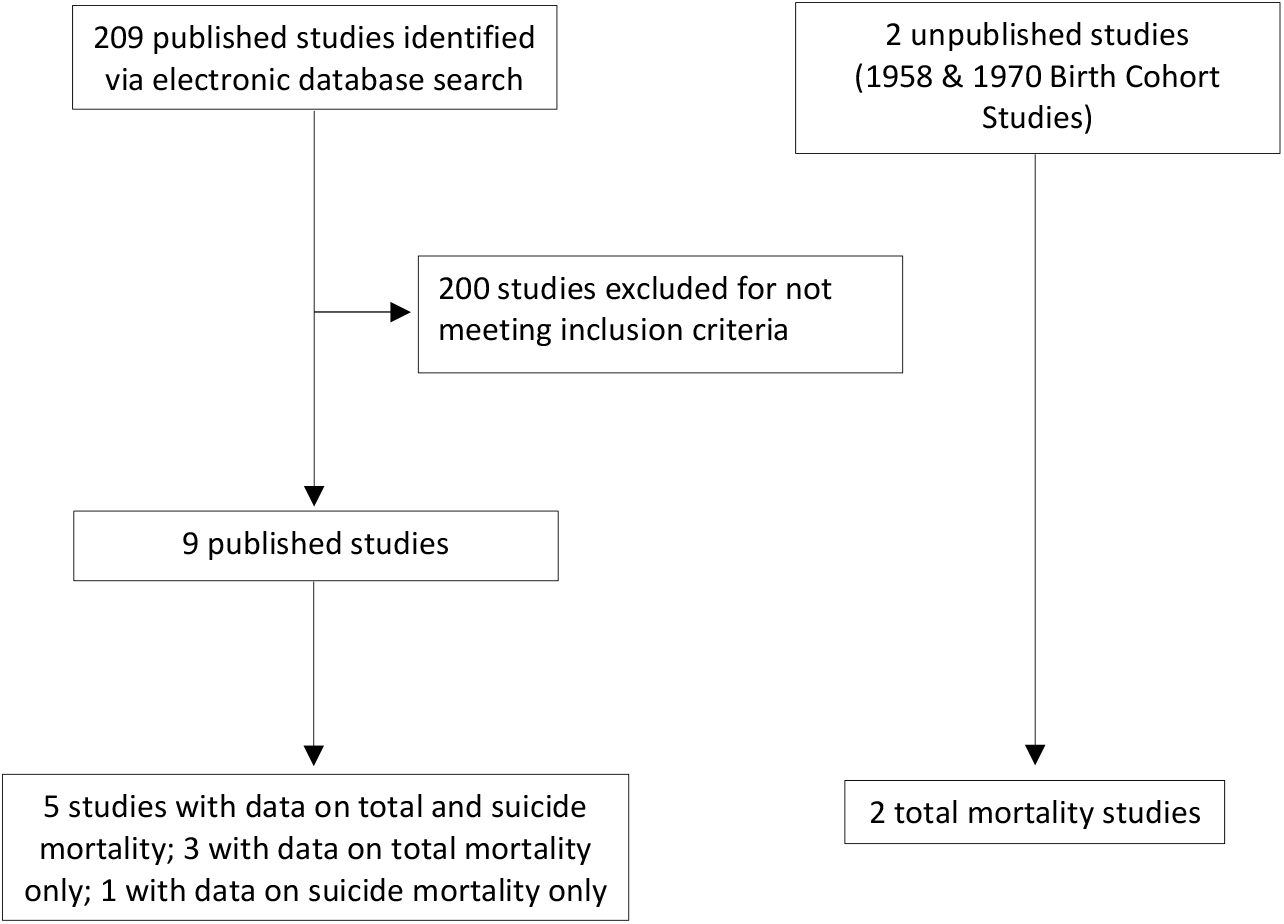
Flow of studies into the review.

Of the ten cohort studies featured in analyses of total mortality, three were drawn from the Swedish population,^10,24,43^ three from the United Kingdom (two unpublished),^45^ and there was one each from the US,^44^ Canada,^42^ Finland,^23^ and Australia^26^ (**table 1**). These studies comprised 2,273,998 individuals (sample size range: 1242^44^ to 989871^43^) born over a 50 year period between 1953^10,45^ and 2003;^26^ the maximum age at follow-up was 65 years.^10^ Three studies used the general population as an external comparator group,^23,24,42^ while the remaining five featured internal comparators where children were exclusively unexposed to state care.^10,26,43-45^ Six studies were generated solely from linkage of participants to population registers,^10,23,24,26,42,43^ and four were field-based such that care status was reported by parent or study member (two published,^44,45^ two unpublished). Where the number of people in the study sample with a history of care during childhood was reported relative to the population at risk, the prevalence ranged between 1%^43^ and 9%.^10^

**Table 1.** Characteristics of included studies.

| **Study<sup>citation</sup> (country, publication year)** | **Year of birth** | **Number of participants (% women)** | **Age at baseline (years)** | **Age at follow-up (years)** | **Total deaths (suicides)** |
| --- | --- | --- | --- | --- | --- |
| **Unpublished studies** |  |  |  |  |  |
| 1958 Birth Cohort (UK) | 1958 | 17670 (49) | 16 | 16-58 | 1101 |
| 1970 Birth Cohort (UK) | 1970 | 16418 (48) | 16 | 16-43 | 357 |
| **Published studies** |  |  |  |  |  |
| Thompson<sup>42</sup> (Canada, 1995) | 1962-80 | 20471 (NR) | 1-18 | 18-25 | 29 (24) |
| Vinnerljung<sup>24</sup> (Sweden, 2001) | 1969-76 | 13100 (NR) | 1-12 | 19-26 | 103 (26) |
| Kalland<sup>23</sup> (Finland, 2001) | 1973-97 | 13371 (NR) | 0-18 | 18-24 | 68 (35) |
| Hjern<sup>43</sup> (Sweden, 2004) | 1973-82 | 989871 (NR) | 13-17 | 13-27 | 2365 (788) |
| Juon<sup>44</sup> (USA, 2014) | 1960 | 1242 (51) | 7 | 20-44 | 87 |
| Wall-Wieler<sup>25</sup> (Sweden, 2018) | 1973-1980 | 487948 (259275) | 1-18 | 18-40 | (404) |
| Jackisch<sup>10</sup> (Sweden, 2019) | 1953 | 14004 (49) | 0-18 | 19-65 | 1354 |
| Murray<sup>45</sup> (UK, 2020) | 1953-2001 | 353601 (49) | 1-18 | 18-60 | 8814 |
| Segal<sup>26</sup> (Australia, 2021) | 1986-2003 | 331254 (49) | 0-16 | 16-33 | 980 |

NR, not reported.

In **figure 2** we provide study-specific and pooled estimates from unpublished and published studies for the association between care in childhood and adult risk of total mortality; results are minimally adjusted. While heterogeneity was high across individual studies (I^2^=92%, p-value 0.001) – study-specific rate ratios varied between 1.53 and 5.77 – all risk ratios were to the right of the vertical line denoting unity, indicating elevated rates of mortality in adults who had been exposed to care earlier in life. Pooling of effects estimates revealed that children who were exposed to care experienced around two and half times the risk of mortality relative to those without such a history (rate ratio; 95% confidence interval: 2.58; 1.96, 3.39).

**Figure 2.**
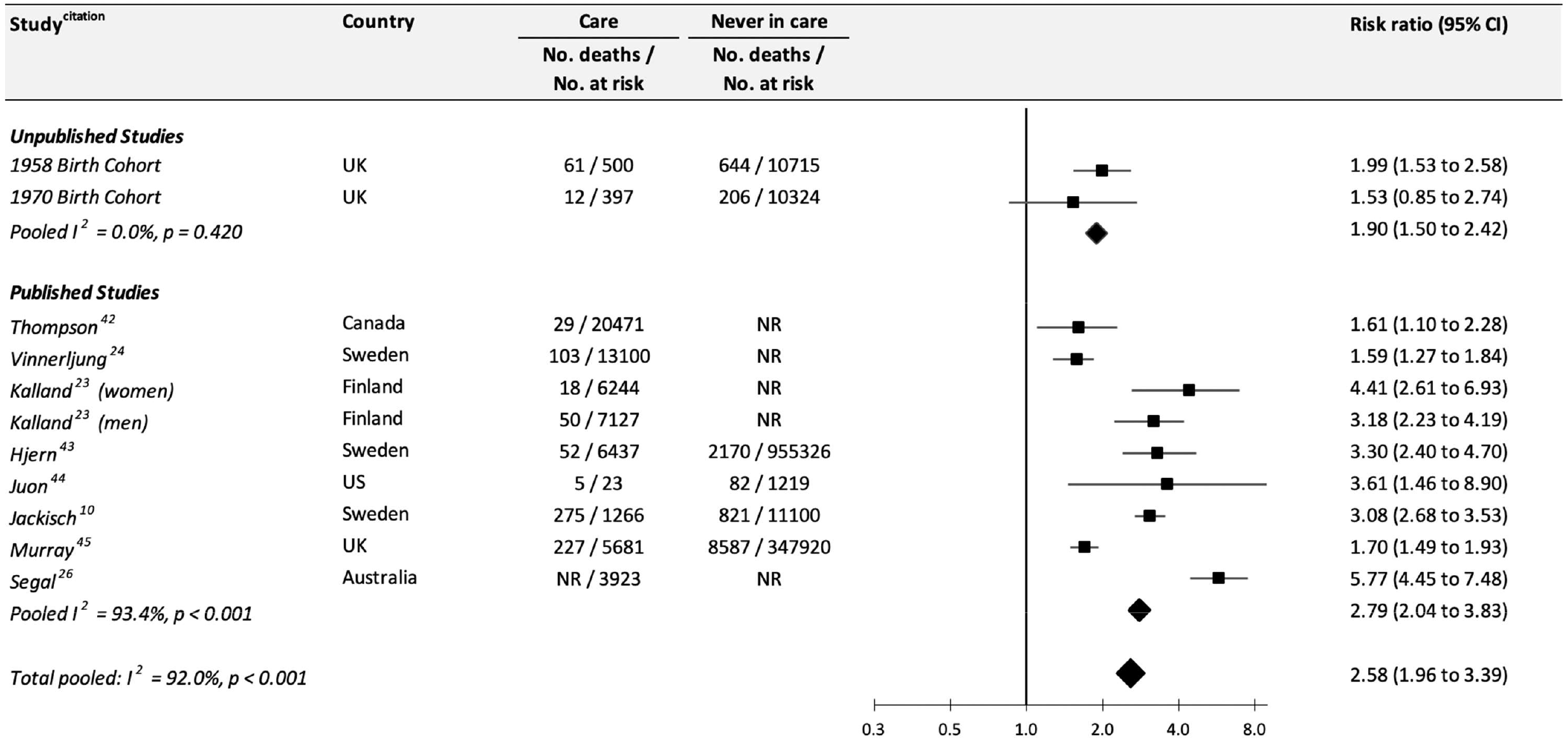
Association between public care in childhood and total mortality by older-age: Meta-analysis of published and unpublished studies. Risk rations are minimally adjusted

Next, we show the association between public care and total mortality according to a different contexts (**figure 3**). Mortality rates were similarly elevated: in both men (2.19; 1.56 to 3.08) and women (2.13; 1.30 to 3.51) who had pre-adult experience of state care; according to duration of follow-up, such that there was around two and a half times the risk of death in individuals followed for up to 6 decades (2.61; 1.74 to 3.94) as there was for early adulthood (2.54; 1.96 to 3.81); and by region, whereby studies initiated in Sweden (2.51; 1.55 to 4.07), the country from which most published data have originated and one which has well-established welfare systems, showed similar effects to elsewhere (2.62; 1.78 to 3.86). There were, however, weaker care–mortality association seen in studies where care history was self-reported either by the respondent or parent (1.80; 1.54 to 2.12) as opposed to being extracted from national registries (2.96; 2.07 to 4.23), and in those studies where investigators used the general population as the external comparison group (2.38; 1.50 to 3.78) rather than a genuinely unexposed internal comparator (2.71; 1.88 to 3.90). Related, the magnitude of the risk ratio for care was lower for those studies evaluated as being of lower quality (2.33; 1.73 to 3.13) versus those that were deemed to be methodologically more rigorous (2.91; 1.90 to 4.45).

**Figure 3.**
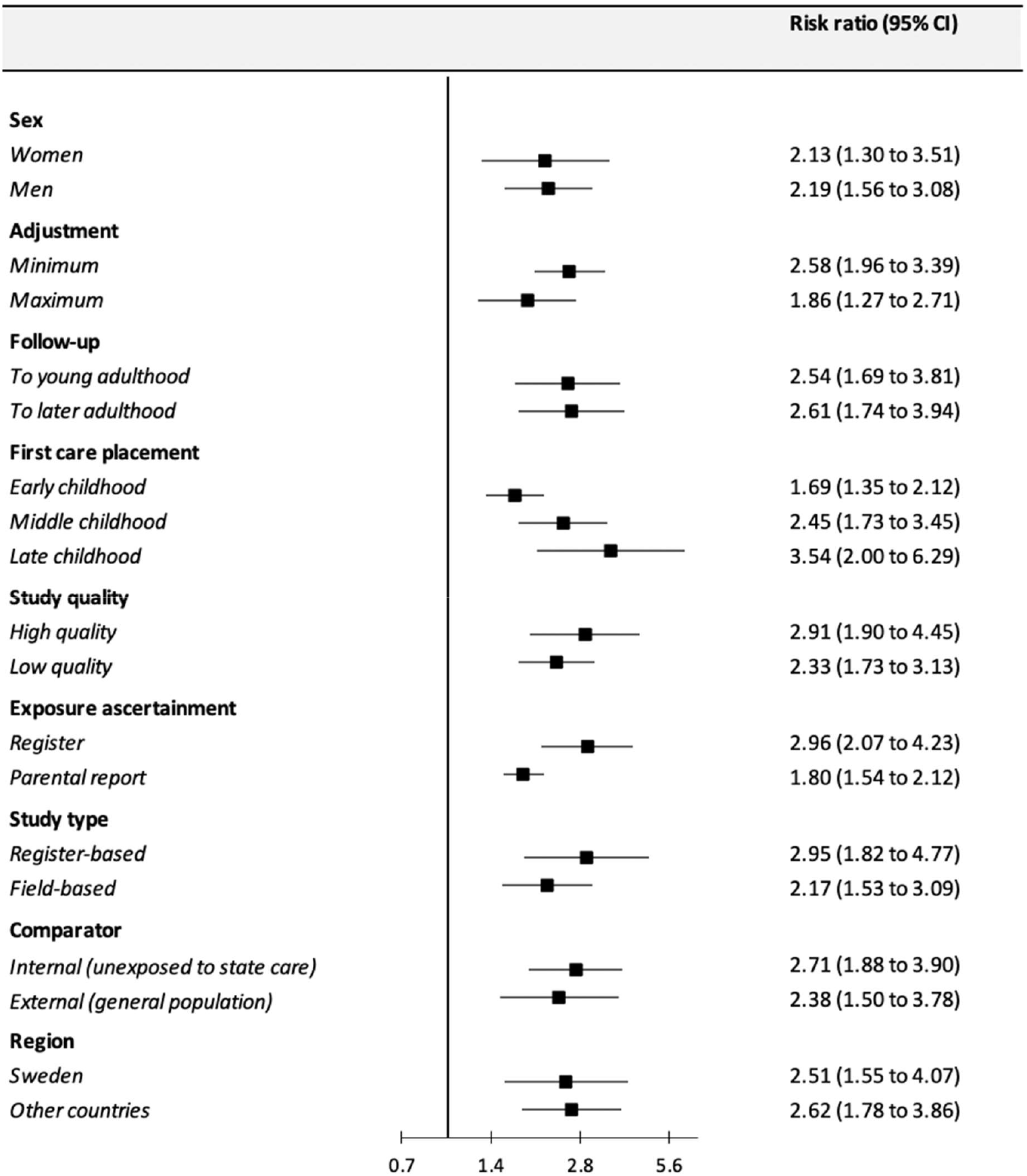
Association between public care in childhood and total mortality by older-age: Analyses according to different contexts. For follow-up, young adulthood is denoted by 18-31 years, and later adulthood by up to age 65 years. For first care placement, early childhood is denoted by ages 0 to 6 years, middle childhood to ages 7 to 12 years, and late childhood ages 13 to 19 years. NR, not reported. For number of deaths and people at risk refer to supplemental figures.

The magnitude of the care–mortality relation was greatest among individuals who had been placed in care in late childhood/adolescence (3.54; 2.00, 6.29) and weaker though still evident among participants entering care in early childhood (1.69; 1.35, 2.12), while children in the intermediate group experienced an intermediate risk of death (2.45; 1.73 to 3.45). Study-specific results according to each of the nine characteristics depicted in **figure 3** are presented supplemental figures 1-9.

Also as demonstrated in **figure 3**, those studies that controlled for a greater array of early life covariates showed markedly weaker relationships (1.86; 1.27 to 2.71) between childhood care and adult mortality than those offering more basic adjustment (2.58; 1.96 to 3.39). With five^23,24,42-44^ of the eight published reports on care and total mortality reporting only rudimentary statistical control (largely age and sex), we used the 1958 and 1970 birth cohort studies to explore in greater detail the impact of individual and collective control for adverse childhood experiences other than care, childhood disability, and poor mental health (**figure 4**). Relative to a comparator model in which effect estimates were initially adjusted for parental socioeconomic occupational social class, mother’s age at birth, and sex (adjustment for age was made indirectly given all study members were born in the same week), we found that taking into account adversity and distress led to some attenuation of risk in the 1958 birth cohort only, while adjustment for disability had no impact on the rate ratios in either set of analyses. Multiple adjustment for these covariates led to an attenuation of almost 50% in the 1958 study (1.54; 1.17 to 2.03) while the 1970 study (1.46; 0.81 to 2.64) was largely robust to such statistical treatment. In related analyses of the two birth cohorts, we also examined if the strength of the relation between care and later mortality was equivalent to that for individuals who had a disadvantaged socioeconomic background (as indexed by parental manual social class) but no experience of care (supplemental table 2). Although statistical precision was hampered by a lower number of deaths in the 1970 cohort, findings from both studies suggested a stronger association between care and mortality than apparent for early life poverty.

**Figure 4.**
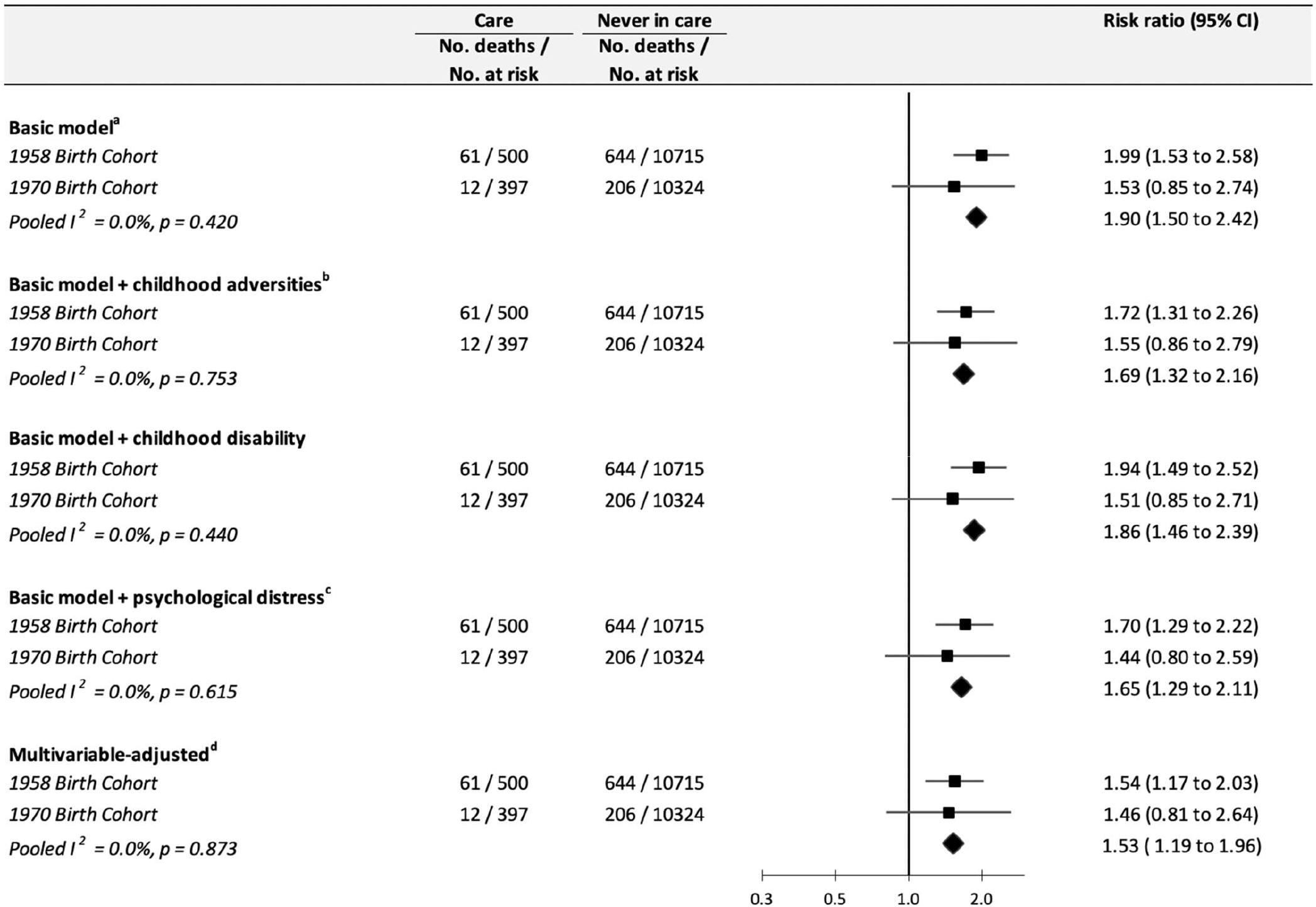
Association between public care in childhood and total mortality by older-age: Meta-analysis of data from unpublished studies with additional statistical adjustments. ^a^sex, parental occupational social class, and mother’s age at birth of participant. ^b^physical neglect; child’s or family’s contact with prison service; parental separation due to divorce, death or, other; family history of mental illness; family history of substance abuse. ^c^psychological distress as measured using the teacher-rated Bristol Social Adjustment Guide (1958 cohort study) and Rutter Behaviour Scale (1970 birth cohort study). ^d^comprises all adjustment

In **figure 5**, we show results for exposure to early life public care and suicide mortality in adult life in five published cohort studies;^23-25,42,43^ cause-specific death data are not currently publicly available in the 1958 and 1970 UK birth cohorts. Three studies were drawn from the Swedish population,^24,25,43^ and one each from Canada^42^ and Finland^23^ (table 1). This material comprised 1,524,761 individuals (sample size range: 13100^24^ and 487,948^25^) born between 1962^42^ and 2003,^26^ the maximum age at follow-up being 39 years.^25^ In all studies, care was associated with higher rates of completed suicide, however, effect estimates were less heterogeneous (*I*^*2*^ = 68%, p-value 0.005) and larger than those apparent in analyses of total mortality, such that the pooled rate ratio suggested a person with experience of care in childhood had a more than three-fold risk of taking their own life in adulthood (3.48; 2.61, 4.64). Study-specific estimates ranged between 2.42 and 5.85.

**Figure 5.**
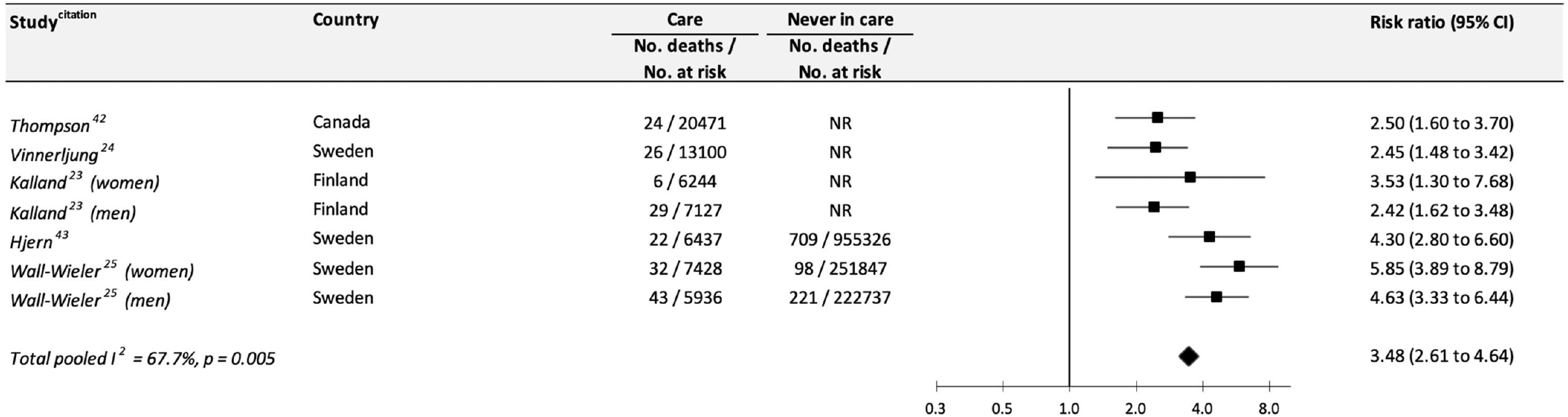
Association between public care in childhood and suicide mortality by middle-age: Meta-analysis of published studies.

With there being fewer suicide studies, opportunities for analyses according to context were diminished, however, as depicted in **figure 6**, the magnitude of the care–suicide association was distinctly weaker in lower quality studies (2.84; 2.25 to 3.60 versus higher quality: 5.08; 3.93 to 6.56), including those using an external comparison group (2.51; 2.00 to 3.15 versus internal: 4.86; 3.90 to 6.06); in men (3.37; 1.79 to 6.37) relative to women (5.34; 3.64 to 7.82); after multiple adjustment for covariates (1.84; 1.16 to 2.91) relative to basic control (4.86; 3.90 to 6.06); and in Sweden (4.13; 2.93 to 5.84) versus other countries (2.54; 1.94 to 3.32).

**Figure 6.**
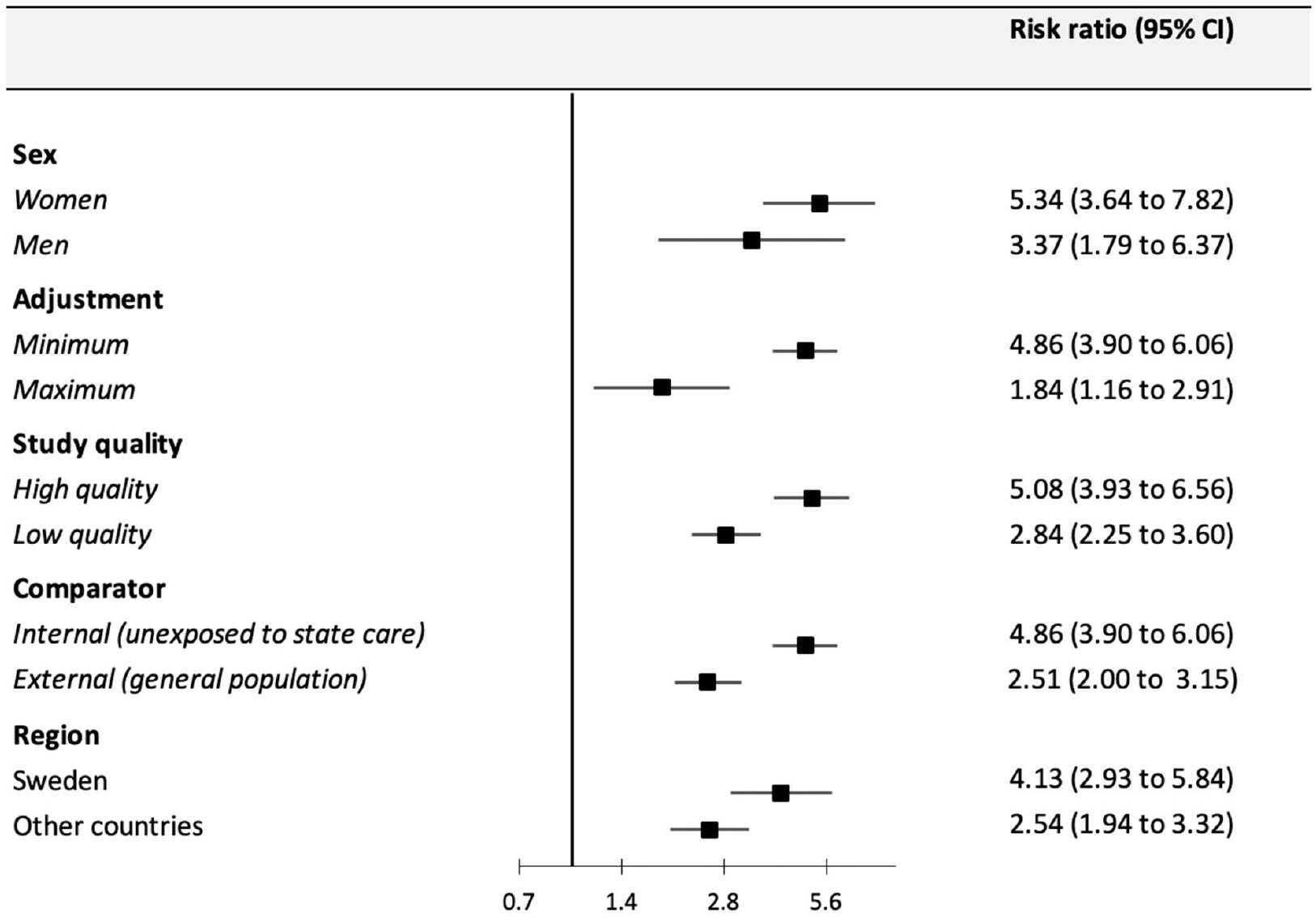
Association between public care in childhood and suicide mortality by middle-age: Analyses according to different contexts.

## Discussion

The main finding of this meta-analysis of published and unpublished prospective cohort studies was that, compared to unexposed children, those with experience of state care had a more than two-fold increased risk of total mortality and three-fold increased risk of suicide in adulthood. The association for care and total mortality, while attenuated, held following statistical adjustment for other early life characteristics known to be related to both entry into care and risk of mortality in later life^38,46^ such as childhood disability, mental health, and other early life adversities (e.g., parental absenteeism or neglect); extended into later adulthood such that it did not exclusively occur immediately following graduation from care; was stronger in better designed studies; and was of equal magnitude in men and women. There was also a suggestion of sensitive periods of exposure to care, whereby individuals who entered public care for the first time in adolescence experienced greater rates of total mortality than those doing so earlier in the life course.

Although based on fewer studies, the magnitude of the association between childhood care and adult risk of completed suicide were higher than for total mortality; similarly, this relationship was not completely explained by control for other early life risk factors in those studies with the capacity to explore confounding. Our overall results for suicide death accord with those from studies of suicide attempt.^47-49^ Given that suicide was often a secondary outcome in retrieved studies owing to the lower number of events, there was typically insufficient reporting by context to facilitate analyses, including for age at follow-up and age at entry to substitute care. There was, however, a suggestion of lower risk in men than women which runs counter to the conjecture that girls may be more resilient to the deleterious impact of care.^50^ The tripling of risk in the suicide analyses suggest that the overall results for total mortality is, in part, being generated by a stronger effect for suicide than for other causes of death in the includes studies, which will include chronic illnesses such as cancer and cardiovascular disease. Too few studies offered results for these other mortality outcomes to facilitate aggregation, however.

There are several potential explanations for our observation from studies of total mortality that adolescence is a sensitive period for placement into care. First, the higher mortality risk apparent for adolescence care may, in fact, be attributed to prolonged exposure to an unfavourable home environment. However, that several of the published studies controlled for childhood socioeconomic circumstances^10,26,45^ and our own analyses of the two birth cohort studies took into account an array of early life adversities (e.g., physical neglect, family history of mental illness/substance abuse) would tend to suggest that there is some risk specifically due to the care experience. It is also the case that removal from care is not universally due to childhood experience of neglect, rather, substitute care may be required because the biological parents are unable to cope with a child’s anti-social behaviour. Second, brain imaging studies have revealed that there is a dramatic acceleration in brain growth during adolescence, with marked development of both cortical and subcortical structures.^52^ Perception of precarious personal circumstances may therefore be more acute at these older ages relative to, for instance, an individual being moved into care earlier in the life course. Moreover, separation from family of origin may also mean removal from the familiar social environments of school and neighbourhood, a change that may trigger adverse health behaviours such as illicit drug use and mental health problems.

That the gradient for substitute care in relation to both total and suicide mortality did not appear to be explained by measured confounding variables implicates direct and indirect mechanisms. Indirect mechanisms include known links between care and unfavourable levels of later sociodemographic,^22^ behavioural,^20^ and health characteristics.^13,16^ A potential direct mechanism is the embodiment of the experience of public care such that there is a biological response to this potentially acute stressor that elevates mortality risk. Mediation by indicators of inflammatory, haemostatic, and metabolic function – known to be associated more broadly with early childhood adversity ^53,54^ – could occur, although in recent analyses of two UK studies there was no suggestion of an association between care and an array of such biomarkers.^6,55^ To further explore mediation by biological factors would require a study with data on care on the one hand, mediators, and mortality outcomes on the other. While we are unaware of such data, as existing cohorts mature, such analyses will become possible. In further analyses of data from the Stockholm Birth Cohort Study – material featured in the present review^56^ – socio-economic and mental health factors would appear to partially mediate the link between public care and total mortality relationship.^33^ An understanding of the role of other candidate mediators including unhealthy behaviours (smoking, heavy alcohol intake, etc), given their relationships with care^20^, and trajectories of physical diseases and death,^57^ is now required.

While the present report has some strengths including its novelty and the use of unpublished individual-level data to complement the findings from published studies, interpretation of our findings inevitably requires consideration of various limitations. Firstly, a meta-analysis is only as methodologically robust as the studies it comprises, and all results in the present review originate from observational data, so raising concerns about unmeasured or residual confounding. In principle, an ideal approach to circumventing the problem of confounding would be to conduct a randomised controlled trial in which only half of children requiring transfer to a safer environment would be allocated to state care. Clearly unethical, a feasible alternative could be a natural experiment whereby the impact of changes in care policy on mortality, such as legislation to reduce the number of children being placed out of the home, are explored. Secondly, that our meta-analysis featured only studies from western populations means that extrapolation of our findings to other countries, particularly when care policies are nation-specific, is moot. Similar research from low- and middle-income countries would be useful. Thirdly, the characteristics of placement into public care in childhood, including type (foster care versus institution), duration, and stability, have been identified as being potentially important in the context of other adverse outcomes.^58^ Results for these characteristics of care were not reported with sufficient frequency in the published studies to facilitate meta-analysis, nor were they collected prospectively in the unpublished datasets. Fourthly, childhood protection and public care policies in several of the countries featured in this review have evolved in recent years to provide better protection for this vulnerable population; as such, historical policies differ to contemporary practice. This notwithstanding, the long-term impact of care policies from many decades ago on people currently in middle- and older-age remains important. Related, short term surveillance of contemporary children with care exposure continues to suggest multiple unfavourable outcomes that are consistent with earlier eras.^59^ Lastly, in studies using a general population external comparison group,^23,24,42^ this would have contained individuals with care experience and, as such, cannot be regarded as being truly unexposed. The lower rate ratios for care and mortality in those studies are likely to be an underestimation of the true effects therefore.

In conclusion, our systematic review and meta-analysis showing excess rates of total and suicide mortality in children exposed to state care from the UK, Sweden, Finland, the USA and Canada suggest child protection systems and social policy following care graduation were, at least in these countries, insufficient to mitigate the effects of the adverse experiences of care itself and the unfavourable events that led to it. These results add to a range of known adverse social, psychological, and behavioural outcomes conventionally thought to occur in early adulthood but, based on this new evidence, seemingly also extending into middle- and older-age.

## Data Availability

Data from the 1958 and 1970 birth cohort studies can be requested from the UK Data Archive (https://www.data-archive.ac.uk/).

## Acknowledgement

We thank participants in the 1958 and 1970 birth cohort studies for their continued forbearance.

---

**Supplemental Table 1.** Assessment of study quality based on the Cochrane Risk of Bias Tool: Studies of public care and total mortality by middle age.

|  | Exposed and unexposed from same population | Confidence in exposure assessment | Comprehensive adjustments | Confidence in confounders assessment | Confidence in outcome assessment | Adequate follow-up | High quality |
| --- | --- | --- | --- | --- | --- | --- | --- |
| **Unpublished studies** |  |  |  |  |  |  |  |
| 1958 study | ++ | + | ++ | ++ | ++ | ++ | Yes |
| 1970 study | ++ | + | ++ | ++ | ++ | ++ | Yes |
| **Published studies** |  |  |  |  |  |  |  |
| Thompson<sup>42</sup> | + | ++ | -- | ++ | ++ | + | No |
| Vinnerljung<sup>24</sup> | + | ++ | -- | ++ | ++ | + | No |
| Kalland<sup>23</sup> | + | ++ | -- | ++ | ++ | + | No |
| Hjern<sup>43</sup> | ++ | ++ | -- | ++ | ++ | + | No |
| Juon<sup>44</sup> | ++ | + | + | + | ++ | ++ | Yes |
| Wall-Wieler<sup>25</sup> | ++ | ++ | ++ | ++ | ++ | ++ | Yes |
| Jackisch<sup>10</sup> | ++ | ++ | + | ++ | ++ | ++ | Yes |
| Murray<sup>45</sup> | ++ | + | - | + | ++ | ++ | No |
| Segal<sup>26</sup> | ++ | ++ | + | + | ++ | + | Yes |

To assess study quality, we responded to the following enquiries: 1. Was selection of exposed and non-exposed cohorts drawn from the same population? 2. Can we be confident in the assessment of exposure? 3. Did the statistical analysis adjust for the confounding variables? 4. Can we be confident in the assessment of the presence or absence of confounding factors? 5. Can we be confident in the assessment of outcome? 6. Was the follow up of cohorts adequate? The studies were evaluated in relation to each question using 4 categories: “definitely yes”, “probably/mostly yes”, “probably/mostly no”, and “definitely no”. The quality of the study was considered high if the responses to each question fell into one of the first two categories.

**Supplemental Table 2.**
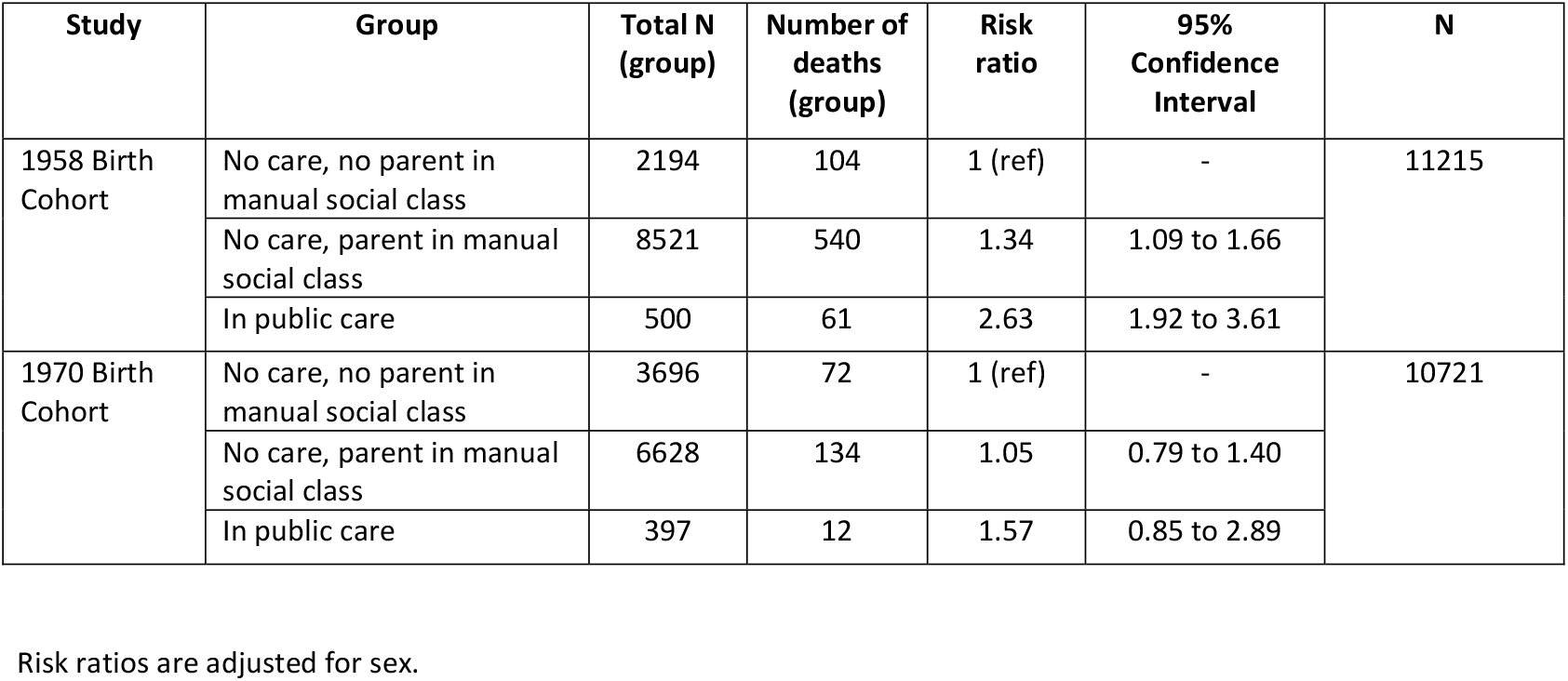
Association of childhood care and early life poverty with total mortality by older-age: The 1958 and 1970 birth cohort studies.

**Supplemental figure 1.**
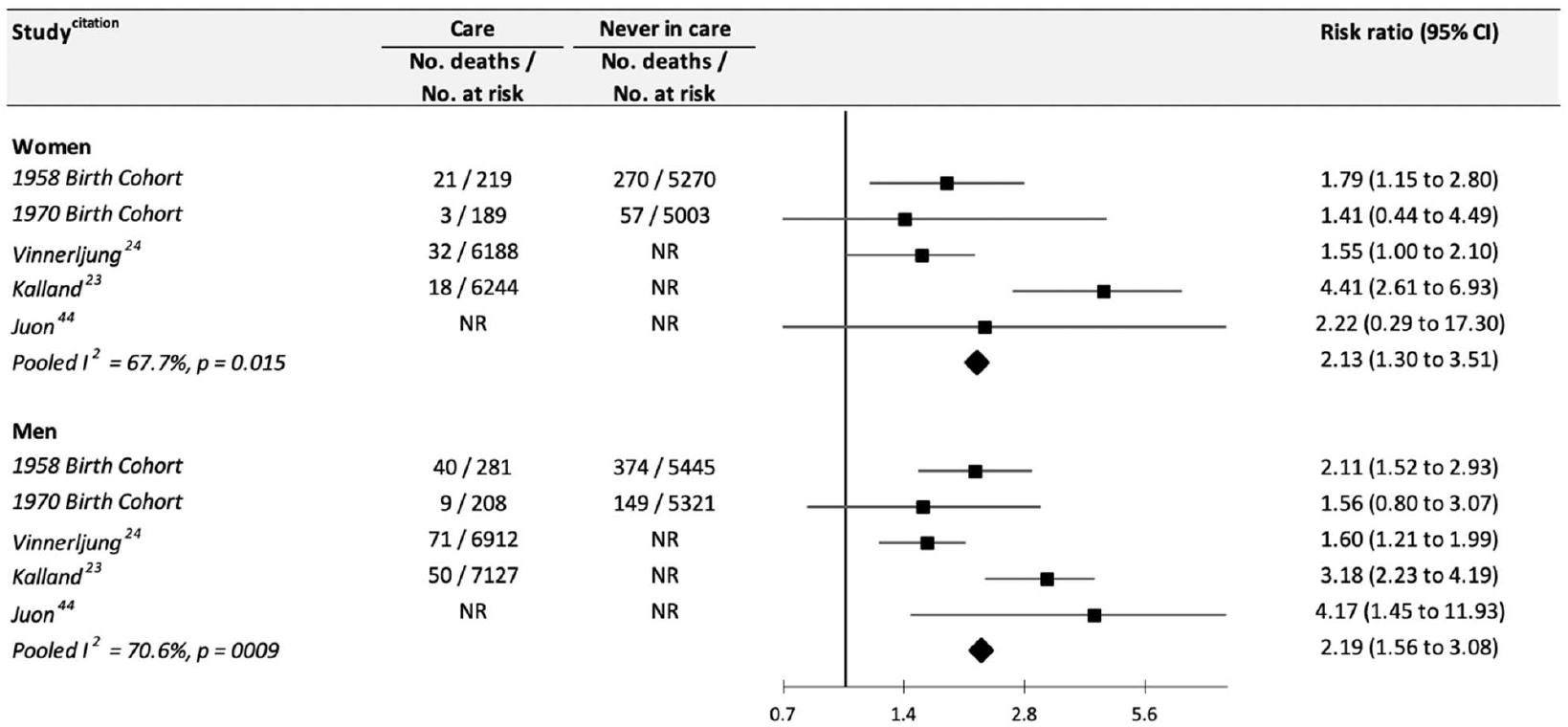
Association between public care in childhood and total mortality by older-age: Study-specific analyses according to sex. NR, not reported

**Supplemental figure 2.**
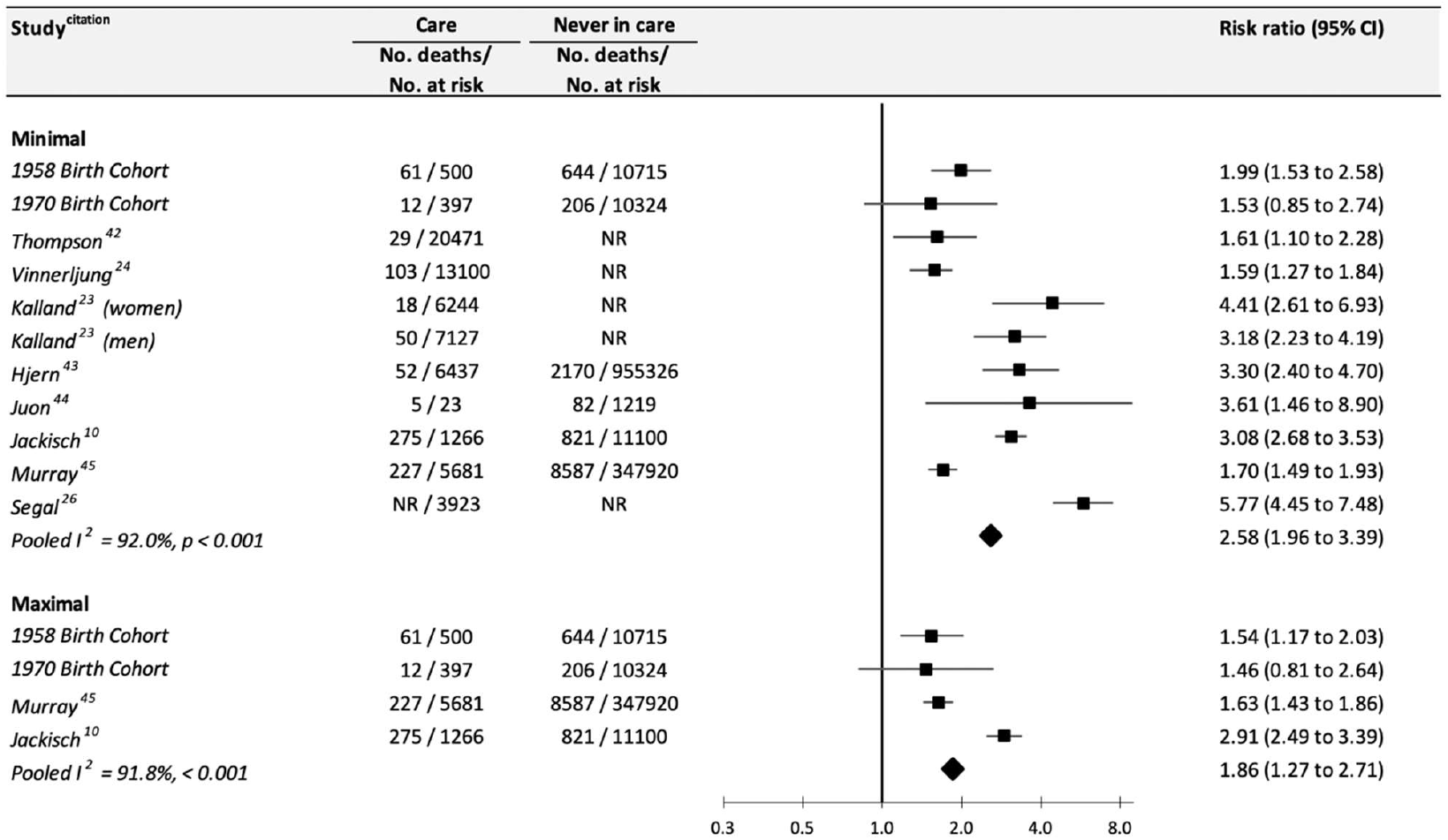
Association between public care in childhood and total mortality by older-age: Study-specific analyses according to level of statistical adjustment. NR, not reported

**Supplemental Figure 3.**
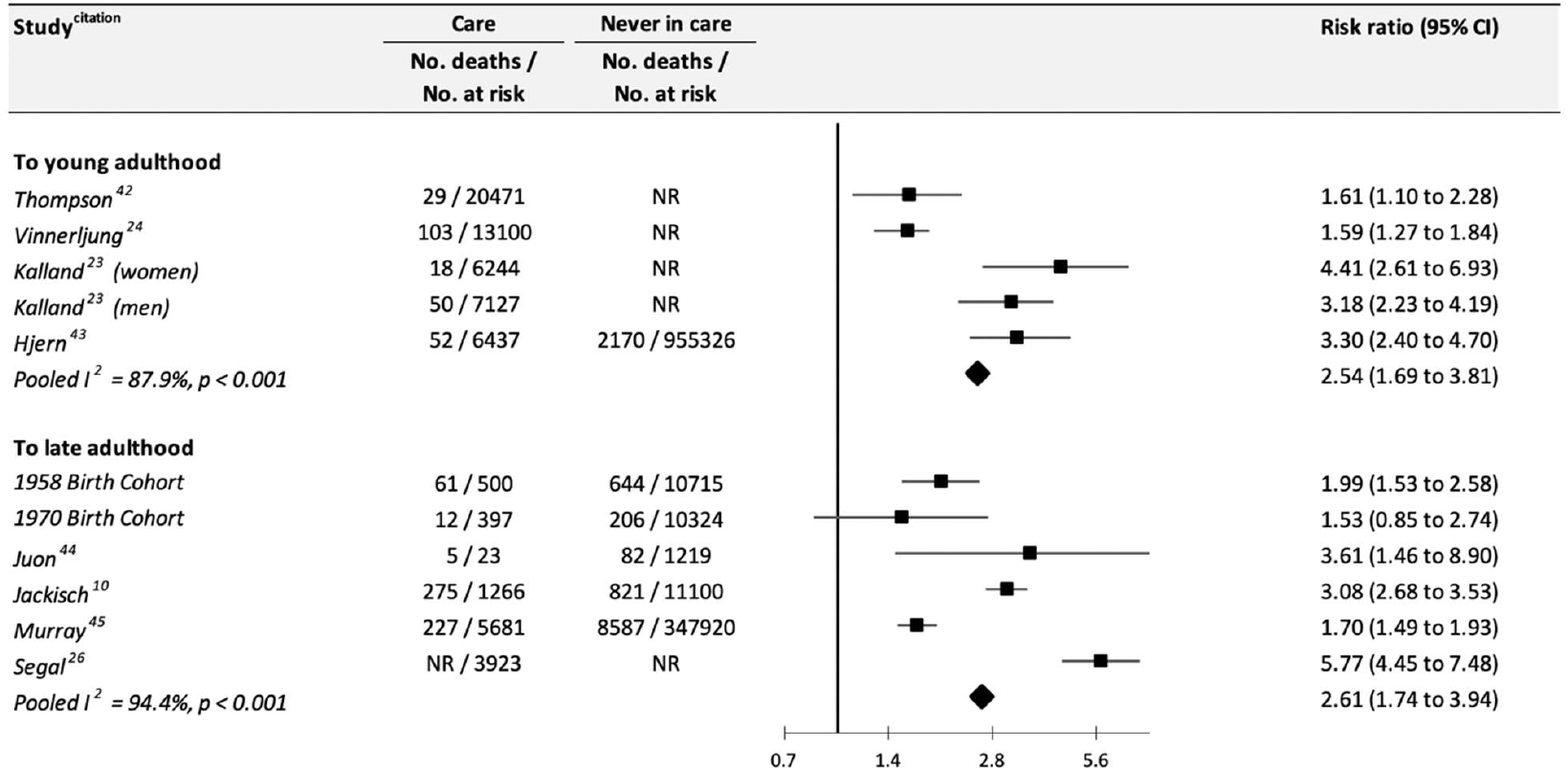
Association between public care in childhood and total mortality by older-age: Study-specific analyses according to duration of follow-up. Young adulthood refers to 18-31 years; later adulthood up to age 65 years. NR, not reported

**Supplemental Figure 4.**
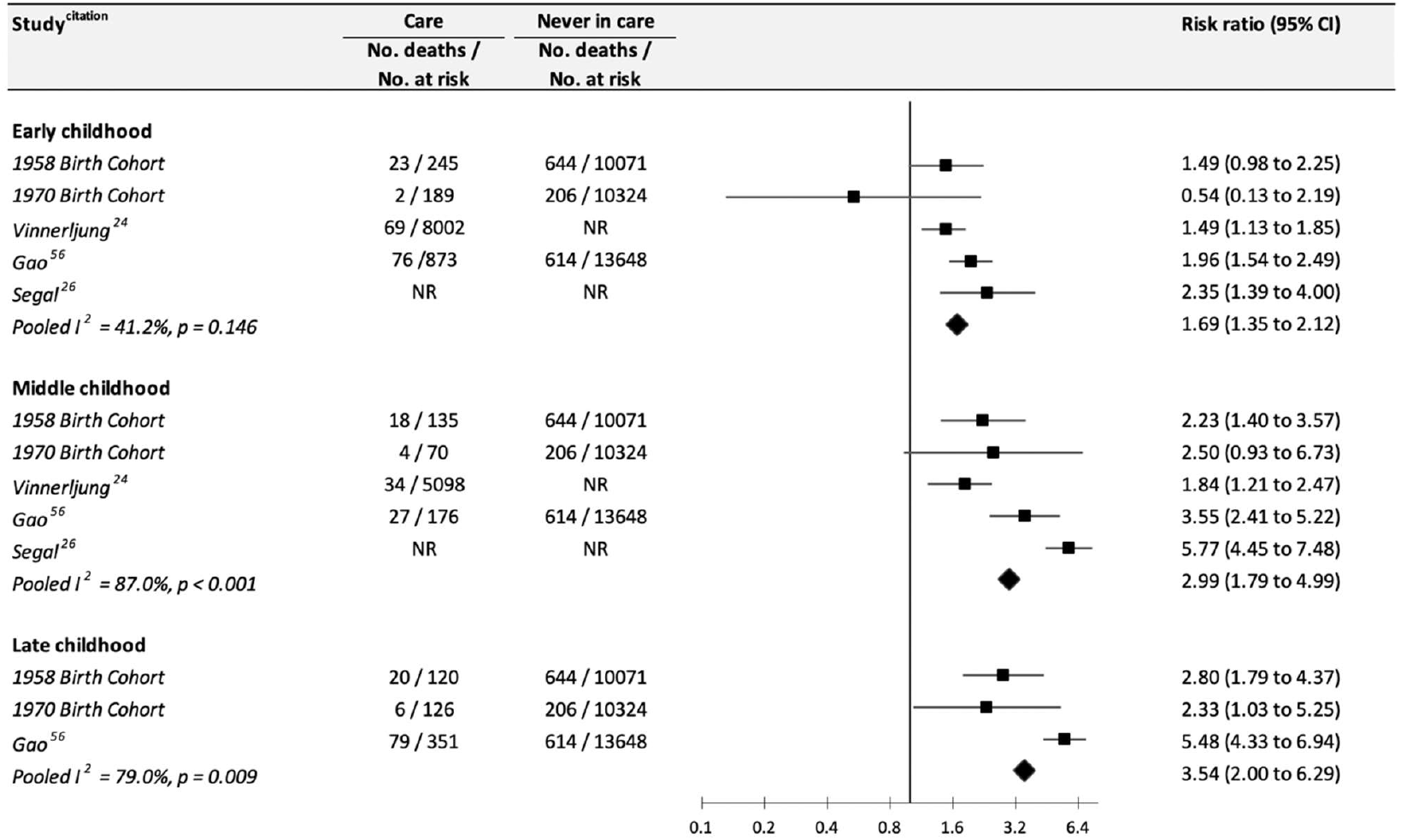
Association between public care in childhood and total mortality by older-age: Study-specific analyses according to age at care placement. Early childhood is denoted by ages 0 to 6 years, middle childhood to ages 7 to 12 years, and late childhood corresponds to ages 13 to 19 years.

**Supplemental figure 5.**
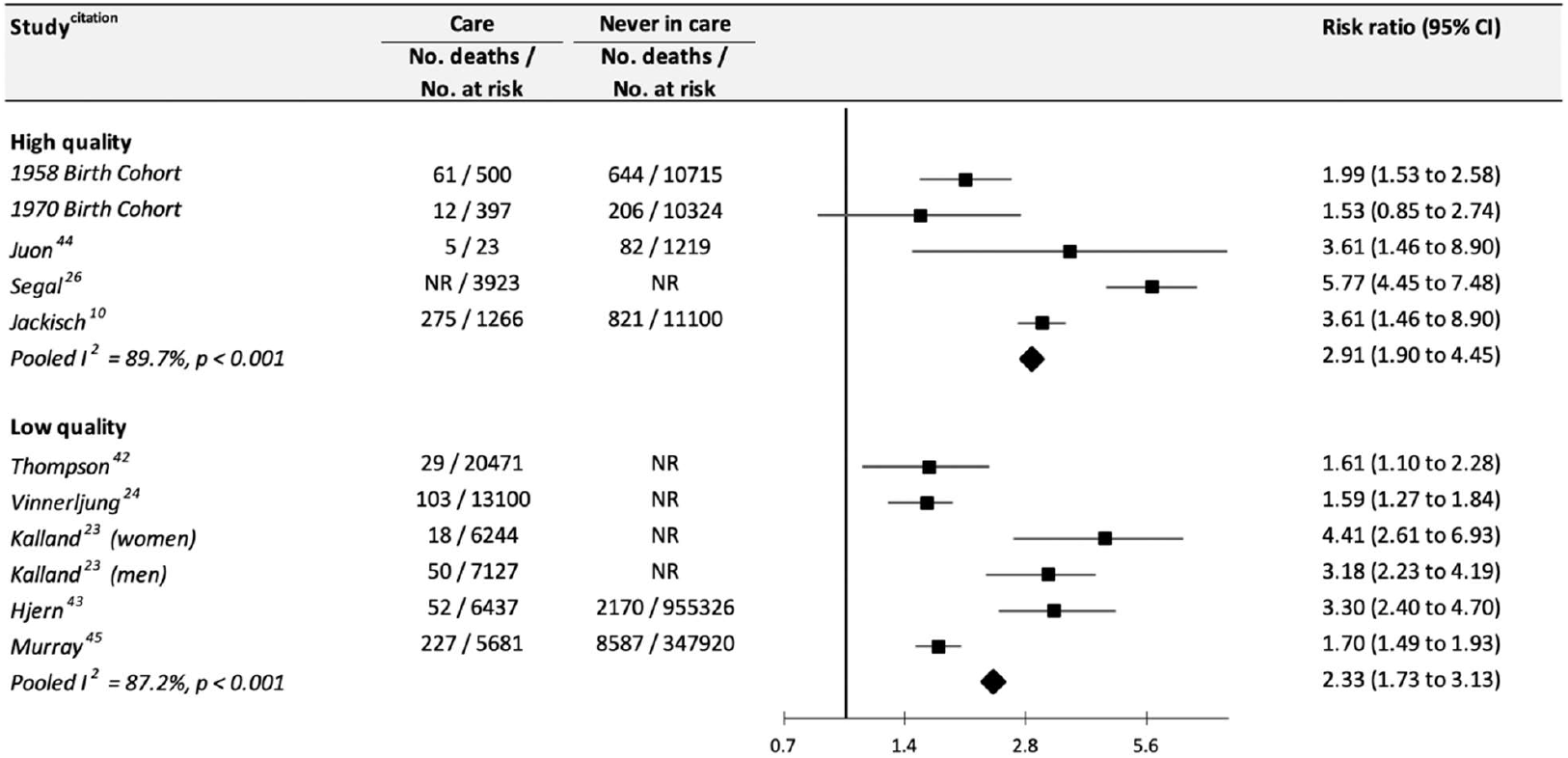
Association between public care in childhood and total mortality by older-age: Study-specific analyses according to study quality. See Supplemental table 1 for study quality criteria. NR, not reported

**Supplemental figure 6.**
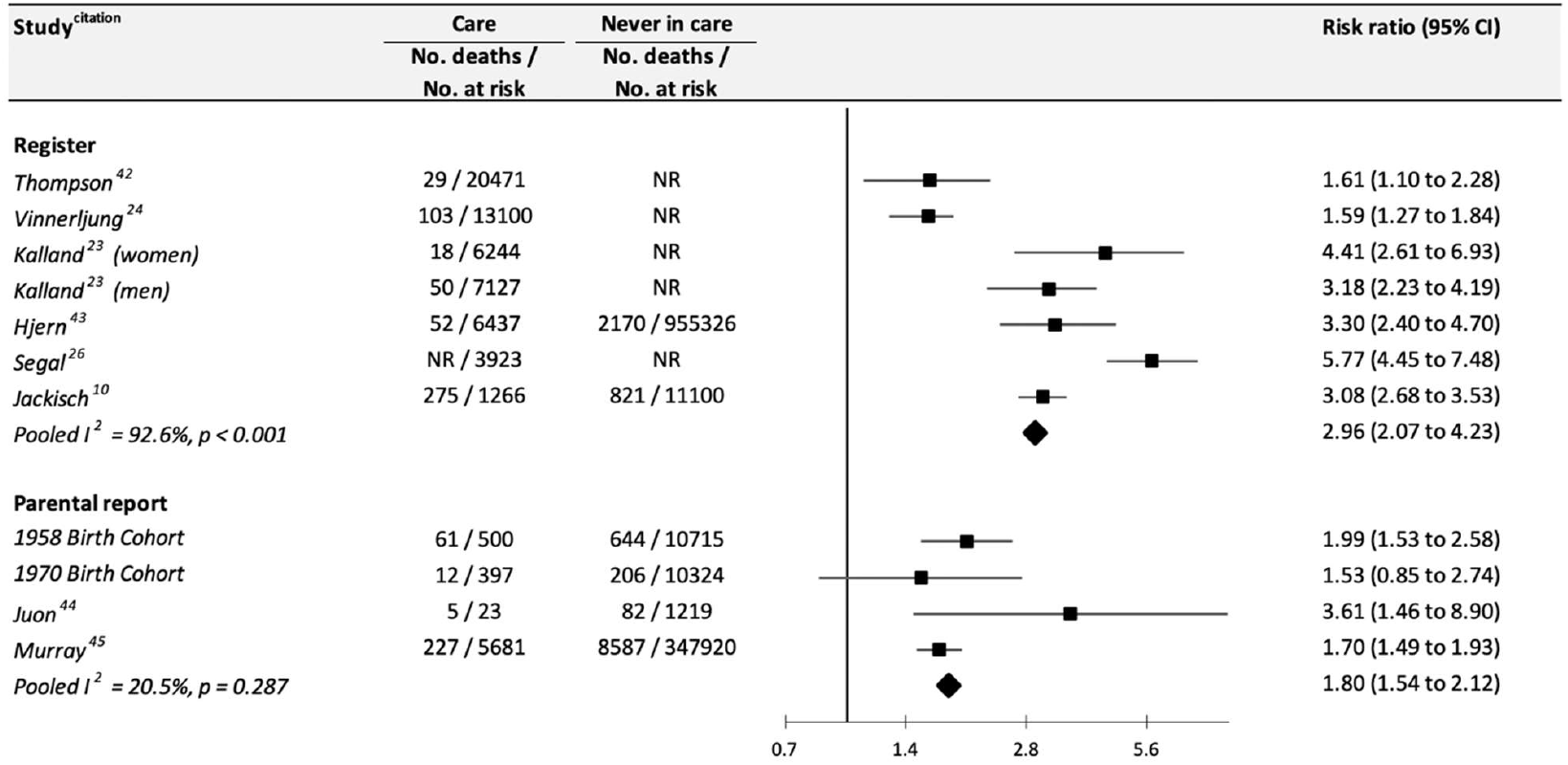
Association between public care in childhood and total mortality by older-age: Study-specific analyses according to method of exposure ascertainment. NR, not reported

**Supplemental figure 7.**
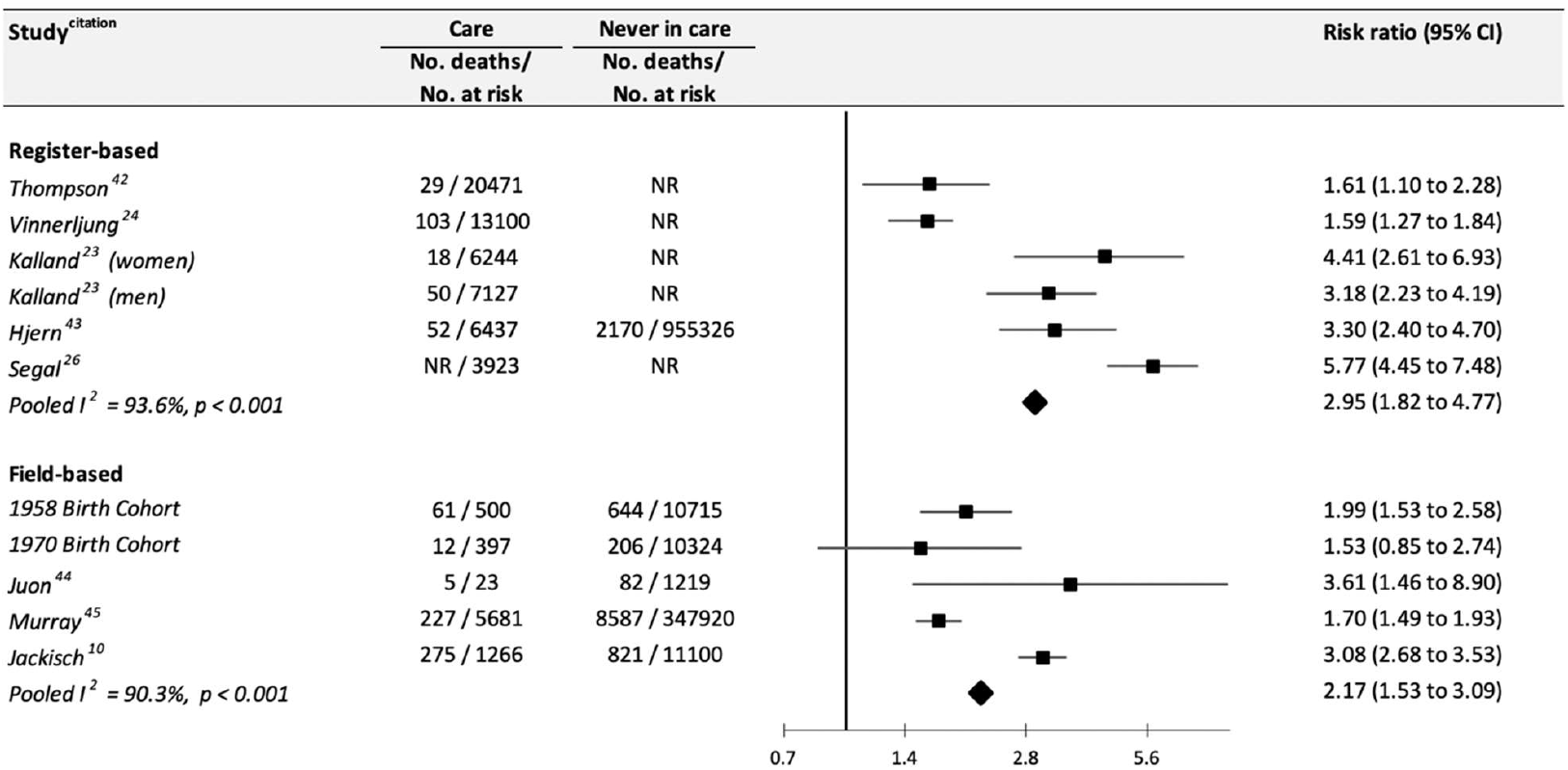
Association between public care in childhood and total mortality by older- age: Study-specific analyses according to study type. NR, not reported

**Supplemental figure 8.**
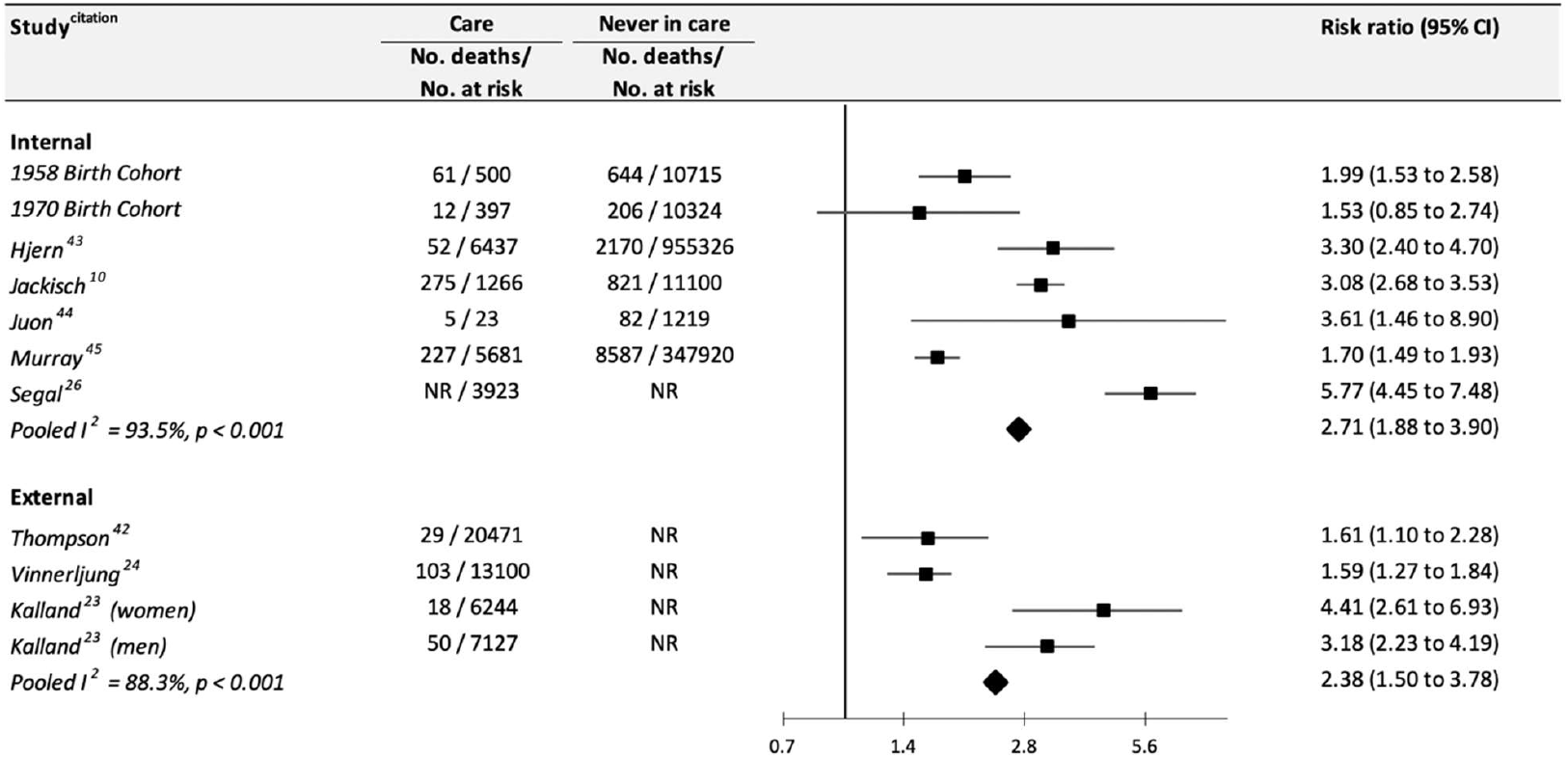
Association between public care in childhood and total mortality by older-age: Study-specific analyses according to exposure comparator group. NR, not reported

**Supplemental figure 9.**
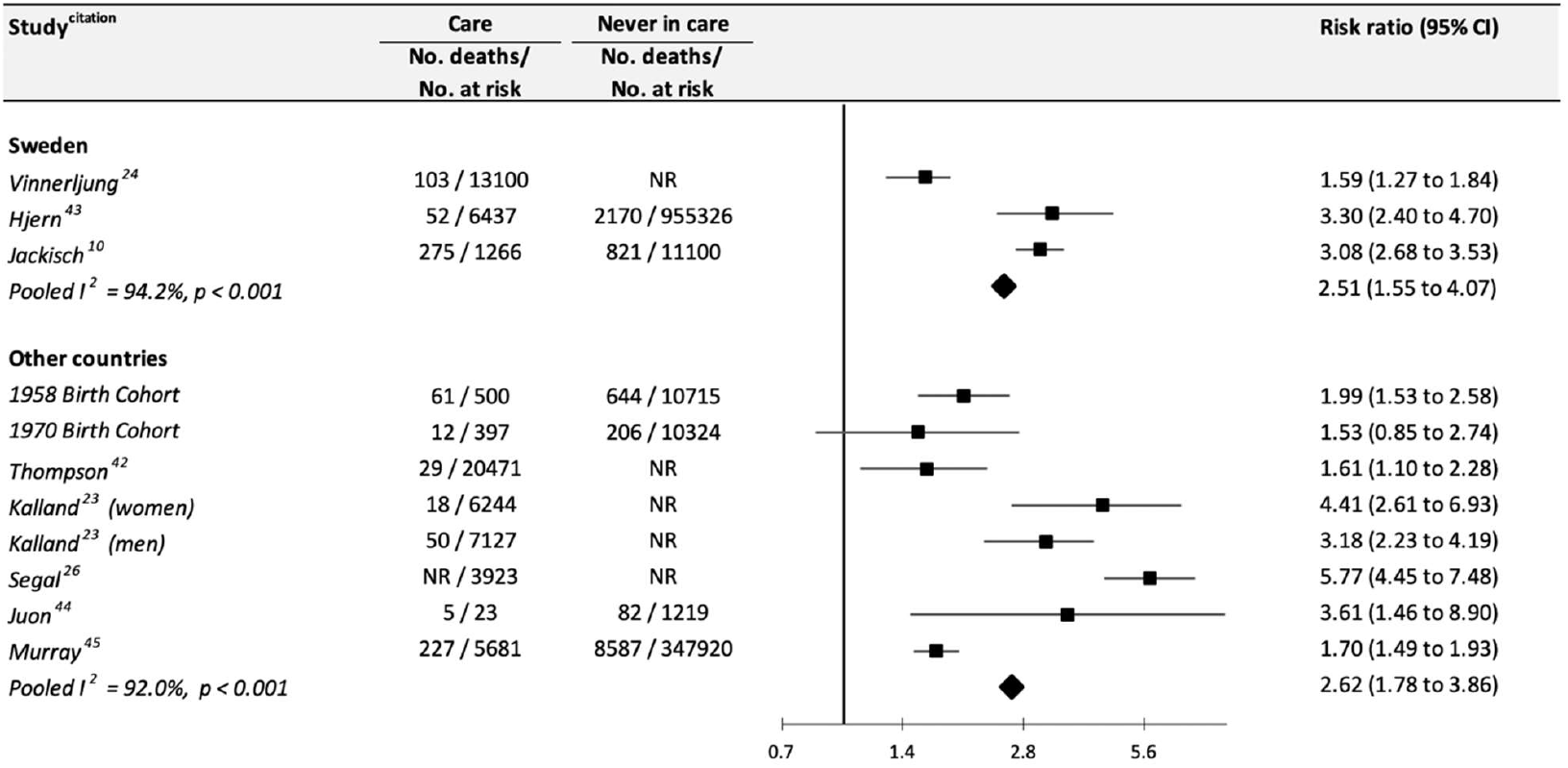
Association between public care in childhood and total mortality by older-age: Study-specific analyses according to region. NR, not reported

## Notes

Conflict of interest: The authors have no conflict of interest.

Funding: GDB is supported by the UK Medical Research Council (MR/P023444/1) and the US National Institute on Aging (1R56AG052519-01; 1R01AG052519-01A1); MK by the Wellcome Trust (221854/Z/20/Z), the MRC (R024227, S011676), NIA (R01AG056477), the Academy of Finland (329202), and Helsinki Institute of Life Science (H970); and PF by the UK Economic and Social Research Council & Biotechnology and Biological Sciences Research Council (Soc-B Centre for Doctoral Training).

### Competing Interest Statement

The authors have declared no competing interest.

### Funding Statement

GDB is supported by the UK Medical Research Council (MR/P023444/1) and the US National Institute on Aging (1R56AG052519-01; 1R01AG052519-01A1); MK by the Wellcome Trust (221854/Z/20/Z), the MRC (R024227, S011676), NIA (R01AG056477), the Academy of Finland (329202), and Helsinki Institute of Life Science (H970); and PF by the UK Economic and Social Research Council & Biotechnology and Biological Sciences Research Council (Soc-B Centre for Doctoral Training).

### Author Declarations

Data collection for the 1958 cohort study was approved by the National Health Service Research Ethics committee, and for the 1970 cohort study by the London Central Research Ethics Committee. With present data being anonymised, additional permissions were not required.

